# Radiotherapy combined with EGFR-TKIs for stage III EGFR-Mutated lung cancer: A retrospective cohort study

**DOI:** 10.1101/2024.03.11.24304144

**Authors:** Liu Gang, Wei Yuan, Gan Langge, Gan Mei, Zheng Qingping, Huang Jing

**Author notes:** Correspondence to*: Dr LiuGang, The People’s Hospital of Laibin, No.159, Pangu Avenue East, Laibin, Guangxi 546100, P.R. China.

## Abstract

The present study aimed to evaluate the efficacy and safety of combining thoracic RT with epidermal growth factor receptor (EGFR)-tyrosine kinase inhibitors(TKIs) in managing stage III lung cancer with EGFR mutation. Cases of patients with stage III EGFR-mutant lung cancer who received thoracic RT between December 2014 and December 2022 from multiple hospitals including The People’s Hospital of Laibin, The First People’s Hospital of Yulin and Guangxi Medical University Kaiyuan Langdong Hospital, were collected. The patients were divided into two groups based on the initial treatment approach: RT + TKIs(RT+TKI) group and RT + chemotherapy (RT+CT) group. The primary measure of interest was progression-free survival (PFS), and additional measures evaluated included objective response rate (ORR), overall survival (OS), patterns of treatment failure and adverse events. Survival analysis was performed using the Kaplan-Meier method, and the log-rank test was used to compare survival rates among different subgroups. A total of 54 patients were ultimately included, with 36 in the RT+TKI group and 18 in the RT+CT group. Regarding short-term efficacy, the ORR for the RT+TKI and RT+CT groups were 86.1 and 66.7%, respectively, with no statistically significant difference (P=0.189). Regarding long-term efficacy, the median PFS for the RT+TKI and RT+CT groups was 26.0 and 11.0 months, respectively, showing a significant difference (P<0.01). The 3 and 5-year OS rates between the RT+TKI and RT+CT groups did not exhibit statistical significance (P=0.825). Subgroup analysis revealed a statistically significant difference in PFS between the combination of RT with third-generation TKIs and first-generation TKIs (P=0.046). The Del19 subgroup exhibited a prolonged median PFS compared with the L858R subgroup, although the difference was not statistically significant (P=0.854). In terms of adverse reactions, the incidence rates of grade ≥3 hematological toxicity and gastrointestinal reactions in the RT+TKI group were significantly lower than those in the RT+CT group (P<0.05). However, the incidence rate of grade ≥3 radiation-related pneumonitis was similar between the RT+TKI and RT+CT groups, with no statistically significant difference. The results suggest that combination of RT and TKIs has superior efficacy and is a safer therapeutic approach for stage III EGFR-mutated lung cancer compared with concurrent radio-chemotherapy.

The present study was registered on the ClinicalTrials.gov website on 2nd June 2023, with the trial registration number NCT05934461.

## Introduction

Lung cancer is a prevalent malignancy, ranking second in terms of incidence and first in terms of mortality among all tumor types(1). A distinct subtype of lung cancer is characterized by mutations in epidermal growth factor receptor (EGFR), with a mutation rate of 51.4% observed in lung adenocarcinoma in China and the Asia-Pacific region(2). It has been reported that EGFR-mutated lung cancer is associated with a less favorable prognosis compared with wild-type EGFR tumors(3).

Tyrosine kinase inhibitors (TKIs), a class of small molecule protein kinase inhibitors, have shown efficacy in suppressing tumor growth by competitively binding to EGFR tyrosine kinase and inhibiting EGFR activity. Randomized controlled trials have provided evidence for the effectiveness and safety of EGFR-TKIs in the treatment of stage IV EGFR-sensitive mutated lung cancer(4–7). Consequently, EGFR-TKIs have become the recommended first-line treatment in major oncology guidelines for stage IV EGFR-sensitive mutated lung cancer. However, the optimal treatment strategy for inoperable stage III non-small cell lung cancer (NSCLC) with EGFR mutation remains a challenge. Concurrent chemoradiotherapy remains the standard of care for inoperable NSCLC in stage III with EGFR mutation (8). The RTOG9410 trial reported a dismal 5-year survival rate of 16% for patients with inoperable stage III NSCLC treated with concurrent chemoradiotherapy(9). The high recurrence rate and toxic side effects associated with this treatment approach have led to limited progress.

Preclinical investigations have demonstrated the synergistic potential of combining RT with TKIs(10, 11). TKIs can arrest tumor cells in the G2/M phase, thereby augmenting radiosensitivity and impeding the repair of radiation-induced DNA damage, consequently enhancing the therapeutic efficacy of radiation. Previous clinical studies on the integration of RT with targeted therapy for stage III inoperable lung cancer predominantly focused on combining chemoradiotherapy with either EGFR-TKIs or anti-EGFR monoclonal antibodies. However, these studies did not incorporate genetic sequencing, resulting in suboptimal outcomes(12–17). With the advancements in genetic sequencing technology, the integration of targeted therapy and RT based on genetic sequencing has garnered substantial attention among clinicians. Two studies, namely the multicenter phase II clinical study RECEL9(18) conducted in China and the small-sample study WJOG691110(19) conducted in Japan, have demonstrated the superiority of RT combined with erlotinib or gefitinib over chemoradiotherapy in the treatment of stage III unresectable EGFR-mutated lung cancer. However, large-scale clinical studies comparing the efficacy of RT combined with targeted therapy versus concurrent chemoradiotherapy are still lacking.

The aim of the present study was to retrospectively analyze the clinical data of patients with stage III EGFR-mutated lung cancer who underwent combined RT and TKIs treatment at The People’s Hospital of Laibin, The First People’s Hospital of Yulin and the Guangxi Medical University Kaiyuan Langdong Hospital, in order to investigate the efficacy and safety of combining RT with EGFR-TKIs for the treatment of stage III NSCLC with EGFR mutation, thus providing valuable insights for clinical practice.

## Materials and methods

### Study design and patients

The present study is a multicenter retrospective cohort study that systematically collected data from December 2014 to December 2022 for patients with stage III EGFR-mutant NSCLC undergoing thoracic RT. We obtained the data access date as 25 December, 2022. The participating medical institutions included The People’s Hospital of Laibin, The First People’s Hospital of Yulin and Guangxi Medical University Kaiyuan Langdong Hospital. Information was comprehensively gathered through electronic medical record systems, including patient demographics (age, sex, ethnicity and smoking status), pathological characteristics, imaging details, Tumor-Node-Metastasis (TNM) staging, functional status scores, genetic testing results, RT, chemotherapy, targeted therapy, and post-treatment information on tumor progression.

The following criteria were used for inclusion: i)Pathologically confirmed initial diagnosis of NSCLC; ii)EGFR mutation confirmed by gene sequencing, including the identification of the exon 19 deletion or L858R mutation; iii)clinical stage determined as stage III NSCLC following the International Association for the Study of Lung Cancer 8th edition TNM staging system; iv)treatment involving thoracic RT combined with TKI or thoracic RT combined with chemotherapy; and v)Eastern Cooperative Oncology Group (ECOG) physical status score ≤ 2. The following criteria were used for exclusion: i) Patients who underwent surgery; ⅱ) patients solely receiving targeted therapy; iii) patients with the initial treatment modality of RT+CT+TKI. iv) patients with pathological types such as squamous carcinoma, neuroendocrine carcinoma, sarcoma; v) patients with other serious diseases that could affect survival; and vi)patients with incomplete staging information.

Regarding the uniform of staging, our team has stipulated, during the patient screening process, that included patients must undergo a physical examination, enhanced chest CT, enhanced MRI or CT of the brain, ultrasound or CT of the neck/clavicular lymph nodes, enhanced CT or ultrasound of the upper abdomen, and a whole-body bone scan or PET/CT. Only patients clinically staged as stage III non-small cell lung cancer (NSCLC) were included in the study. Additionally, the accuracy of staging was independently assessed by two senior doctors using the International Eighth Edition of the Lung Cancer TNM Staging System.we conducted brain-enhanced MRI or enhanced CT scans to confirm that patients without brain metastasis before treatment were included.

The enrolled patients were categorized into two groups based on the initial treatment modality: Thoracic RT combined with TKI (RT+TKI) and thoracic RT combined with chemotherapy (RT+CT) groups, with RT+TKI as the study group and RT+CT as the control group.

### Treatment options

#### Study group

(RT+TKI): RT used the three-dimensional conformal RT (3DCRT) or intensity-modulated RT (IMRT) techniques. The delineation of target areas follows the International Commission on Radiation Units and Measurements 83 guidelines. The gross tumor volume (GTV) included visible lesions on CT (including lymph nodes > 1 cm in diameter or pathologically confirmed lesions). The clinical target volume (CTV) encompassed subclinical lesions surrounding the GTV, with an 8 mm outgrowth. The planned target volume (PTV) boundaries were set 5-10 mm outside the CTV boundary, depending on the magnitude of respiratory motion. A prescribed dose of 54-66Gy/27-33 fractions was administered. Dose limits for organs at risk were as follows: Lung V20 < 30%, mean lung dose (Dmean) < 17 Gy, spinal cord Dmax < 45 Gy and heart V30 < 40%.

Regarding targeted therapy: the TKIs used in the study included gefitinib, erlotinib, icotiniband and osimertinib. The administration of TKIs was as follows: Gefitinib 250mg orally once daily; icotinib 125mg orally three times daily; erlotinib 150mg orally once daily; and osimertinib 80mg orally once daily; TKIs were continuously administered during RT until disease progression or intolerability.

#### Control group

((RT+TKI): RT followed the same protocol as in the RT+TKI group. Chemotherapy consisted of platinum-based doublet regimens, including AP, AC, PC and DP, with specific dosages as follows: AP, pemetrexed 500 mg/m2 on day 1 and cisplatin 75 mg/m2 on day 1; AC, pemetrexed 500 mg/m2 on day 1 and carboplatin AUC 5 mg/ml/min on day 1; PC: carboplatin AUC 2 mg/ml/min on day 1 and paclitaxel 135-175 mg/m2 on day 1; and DP: cisplatin 75 mg/m2 on day 1 and docetaxel 75 mg/m2. Each cycle lasted 21-28 days, with a minimum of 2 cycles and a total of 4-6 cycles.

#### Follow-up visits

The first follow-up visit took place 1 month after the completion of RT, either through a telephone call or in the outpatient clinic, to evaluate the patient’s treatment response. Subsequent follow-up visit intervals were 3 months within 2 years and 6 months within 5 years. These visits included assessments of disease progression and survival outcomes after treatment. The follow-up period concluded in June 2023. Missed cases were recorded based on the patient’s last follow-up, which was used as the cutoff for survival analysis.

### Evaluation of efficacy

#### Recent efficacy evaluation

According to the Response Evaluation Criteria in Solid Tumors (RECIST) version 1.1, complete response (CR) is defined as the complete disappearance of lesions lasting > 1 month, while partial response (PR) involves a reduction in lesion volume of >30% lasting 1 month, and progressive disease (PD) is characterized by an increase of ≥20% in the minimum value of the primary lesion during the course of treatment or the appearance of new lesions. The objective response rate (ORR) was calculated based on the combined CR and PR rates.

#### Long-term efficacy assessment

The major outcome measure of the study was progression-free survival (PFS), which is defined as the period from treatment initiation to the first documented PD or mortality. Secondary outcome measures included overall survival (OS) and treatment failure pattern. OS was determined as the duration from treatment initiation to mortality resulting from any reason. Treatment failure pattern included the progression of the primary tumor and emergence of new lesions.

#### Adverse effects

According to the Common Terminology Criteria for Adverse Events version 5.0 released by the USA Department of Health and Human Services, evaluations were conducted for gastrointestinal reactions and hematological toxicity related to radiation, chemotherapy, or targeted therapy, classified on a scale from 0 to IV. Radiation-induced injuries occurring within the first 3 months after the initiation of RT were defined as acute radiation injuries, while those occurring 3 months or later after RT were classified as late radiation injuries.

#### Statistical analysis

Statistical analysis was conducted using SPSS 27.0 software (IBM Corp.). Descriptive statistics were presented as the mean (range) for continuous variables, while frequencies and percentages were used for categorical variables. Survival analyses were performed using the Kaplan-Meier method. Stratified and unstratified log-rank tests were used to compare PFS and OS. Survival curves were generated using GraphPad Prism 9.5 software (GraphPad; Dotmatics). All statistical tests were two-sided. P<0.05 was considered to indicate a statistically significant difference.

## Results

### Baseline Patient Characteristics

Between December 2014 and December 2022, a screening of 3,684 patients with NSCLC patients was conducted. Among them, 1,289 patients underwent genetic testing, revealing 547 cases with EGFR mutations, resulting in a mutation rate of 42.4%. Through further selection, 112 patients with stage III EGFR mutations were identified and included. Exclusions comprised 26 patients who underwent surgical treatment, 25 patients receiving single-targeted therapy, 3 patients with incomplete staging information, 3 patients with severe complications, and 1 patient with squamous cell carcinoma pathology. Ultimately, 54 patients were enrolled, with 36 in the RT+TKI group and 18 in the RT+CT group. The patient screening process is illustrated in Fig 1.

**Fig 1.**
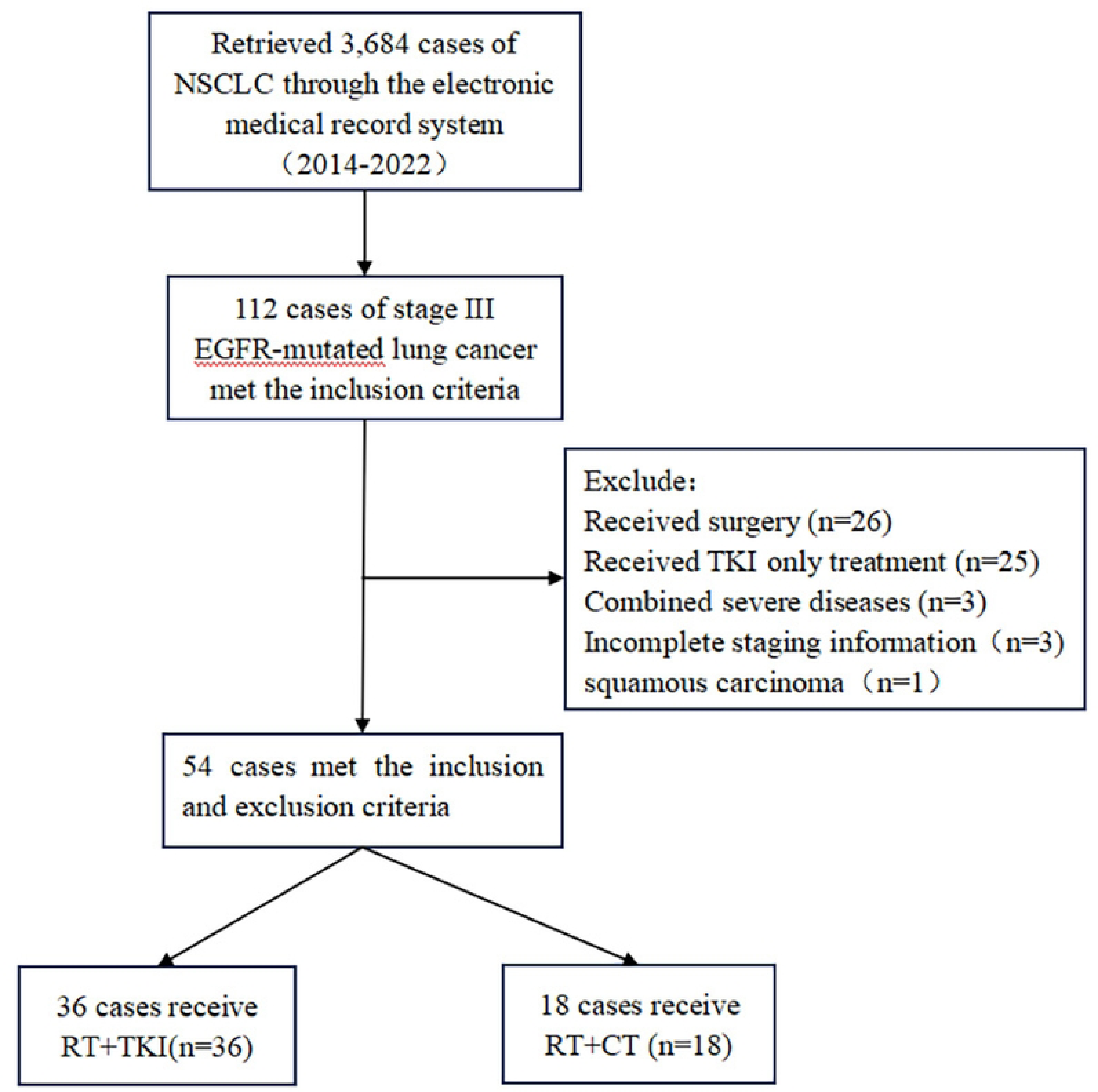
The patient screening process. *Abbreviations:* NSCLC=non-small cell lung cancer; EGFR = epidermal growth factor receptor; TKI=tyrosine kinase inhibitors; RT=radiotherapy; CT=chemotherapy.

Among these 54 patients, 5 underwent PET/CT for staging, while the remaining patients were staged through various examinations, including chest-enhanced CT, cranial-enhanced MRI or CT, neck/clavicular lymph node ultrasound or CT, abdominal-enhanced CT or ultrasound, and whole-body bone scans. These examinations consistently confirmed that all patients had stage III lung cancer (Table I).

### Treatment details

Both the RT+TKI and RT+CT groups employed 3DCRT, IMRT. In the RT+TKI group, the median radiation dose was 60 Gy (range, 54-66 Gy), with a median mean lung dose (MLD) of 14.86 Gy (range: 9.28–16.29 Gy) and a median V20 of 26.3% (range, 23.5-29.9%). In the RT+CT group, the median radiation dose was 60 Gy (range, 56-64 Gy), with a median MLD of 16.08 Gy (range, 10.36-17.20 Gy) and a median V20 of 27.4% (range, 24.61-30.05%).

In the RT+TKI group, the RT combined with TKIs included 17 cases (47.2%) of gefitinib, 8 cases (22.2%) of erlotinib, 2 cases (5.6%) of osimertinib, and 9 cases (25.0%) of afatinib. Among these, 21 cases (58.3%) underwent RT after TKI treatment induction, while 15 cases (41.7%) received concurrent RT and TKI treatment at the beginning. All patients in the RT+TKI group continued to receive TKIs consolidation treatment until tumor progression.

By contrast, the RT+CT group employed RT combined with chemotherapy using AP (10 cases), AC (6 cases) and PC (2 cases). During chemotherapy, 5 cases completed 3 cycles, 8 cases completed 4 cycles, 3 cases completed 5 cycles, and 2 cases completed 6 cycles(Table II).

### Treatment response and suvival outcome

In terms of recent efficacy, the ORR in the RT+TKI group was 86.1%, with 11 cases achieving complete remission (CR), 20 cases achieving partial remission (PR) and 5 cases unavailable for evaluation. The RT+CT group had an ORR of 66.7%, including 2 cases of CR, 10 cases of PR, 4 cases of stable disease (SD), and 2 cases unable to assess efficacy. The difference in ORR between the two groups was not statistically significant (χ2=1.727, P=0.189).(Table III).

In June 2023, there were 2 cases lost to follow-up in the RT+TKI group and 1 case lost to follow-up in the RT+CT group, with a loss to follow-up rate of 5.5%. The median follow-up time was 34.5 months [interquartile range (IQR), 25.8-50.0] for the RT+TKI group and 36.0 months (IQR, 31.0-40.0) for the RT+CT group. The median PFS was 26 months [95% confidence interval (CI), 20.5-31.4] for the RT+TKI group and 11months (95% CI, 8.0-13.9) for the RT+CT group, with a statistically significant difference (χ2=47.088, P<0.01)(Fig 2.). Forest plot analysis revealed that RT+TKI was superior to RT+CT in PFS across subgroups, including age, sex, smoking status, ethnicity, EGFR mutation type, and clinical stage. (Fig 3.)

**Fig 2.**
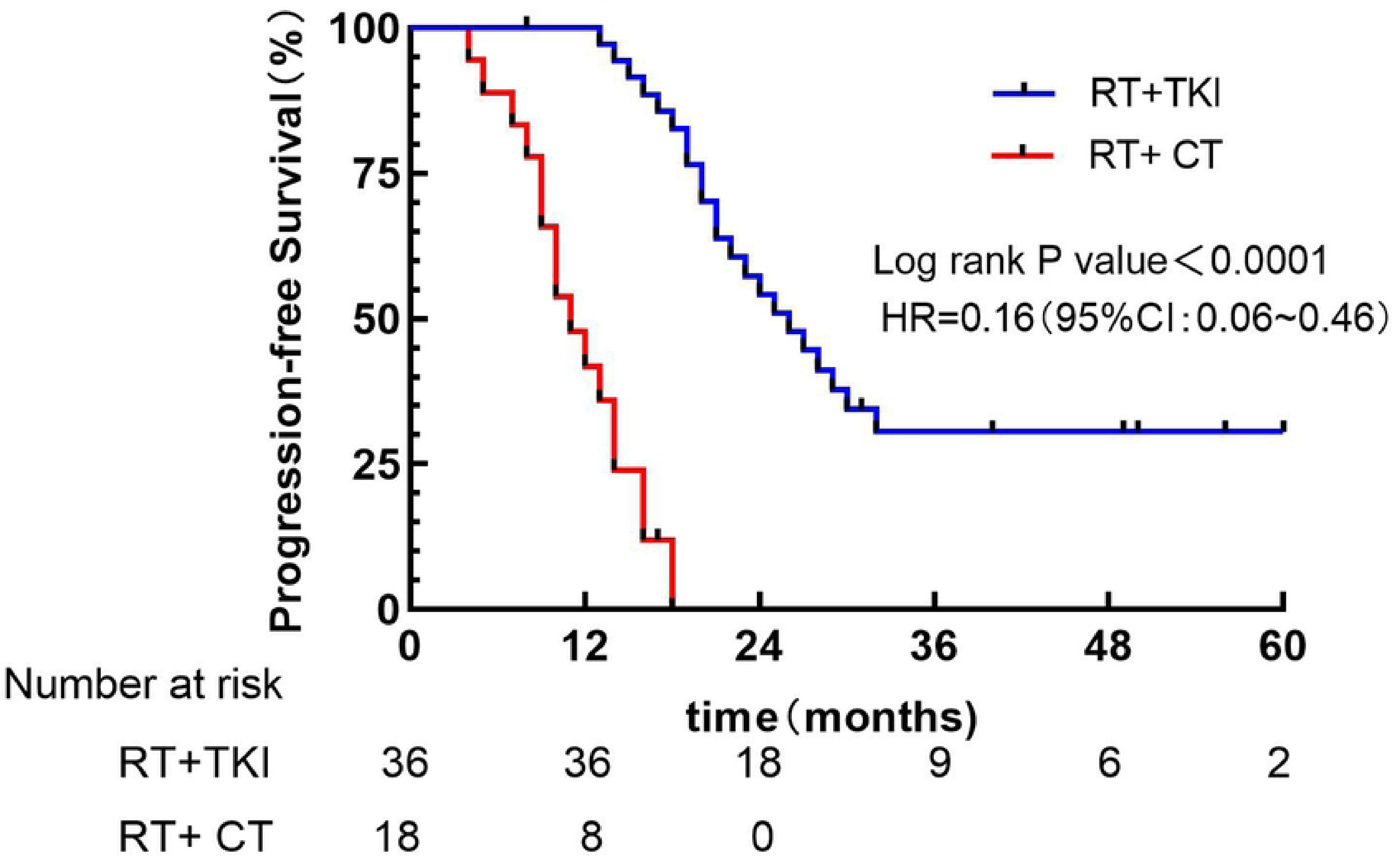
Kaplan-Meier curves of progression-free survival for all patients(N=54). *Abbreviations:* CI=confidenceinterval; HR=hazard ratio; RT=radiotherapy; TKI=tyrosine kinase inhibitors; CT=chemotherapy.

**Fig 3.**
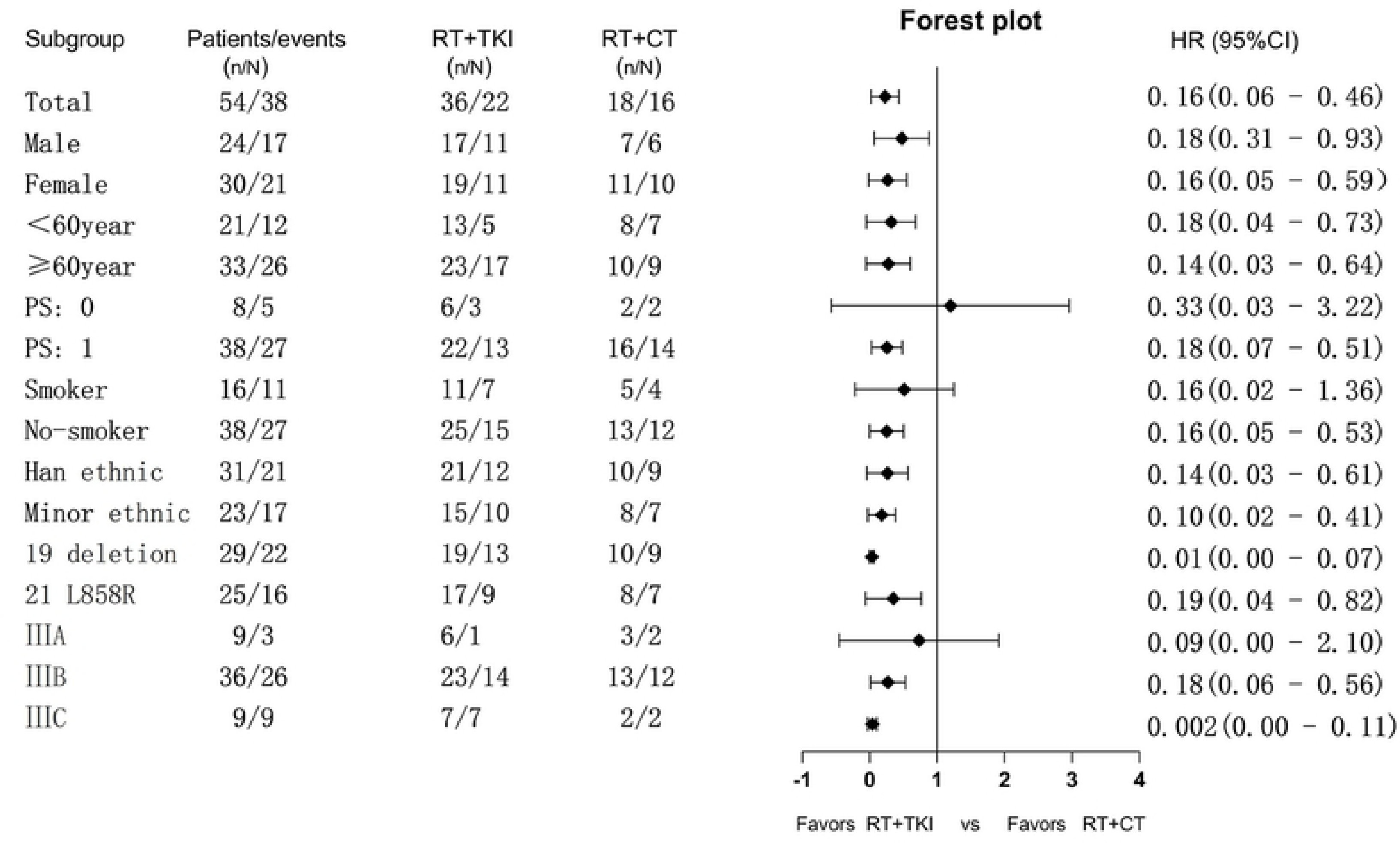
Forest plots for evaluation of treatment effect on different group variable. The analysis was performed on 54 patients with EGFR mutations with PFS as the endpoint. *Abbreviations:* CI=confidenceinterval; HR=hazard ratio; PS=Performance Status; RT=radiotherapy; TKI=tyrosine kinase inhibitors; CT=chemotherapy.

Further analysis of the RT+TKI group showed no statistically significant difference in median PFS between the Del19 and L858R groups (27 vs. 24 months, χ2=0.017, P=0.895)(Fig 4A.). The median PFS for RT combined with third-generation TKI was 32.0 months, and for RT combined with first-generation TKI, it was 22months, with a statistically significant difference (χ2=3.985, P=0.046)(Fig 4B.). The median PFS for the Minority ethnic group was 21 months, and for the Han ethnic group was 28months, with no statistically significant difference (χ2=1.754, P=0.1854)(Fig 4C.).

**Fig 4A.**
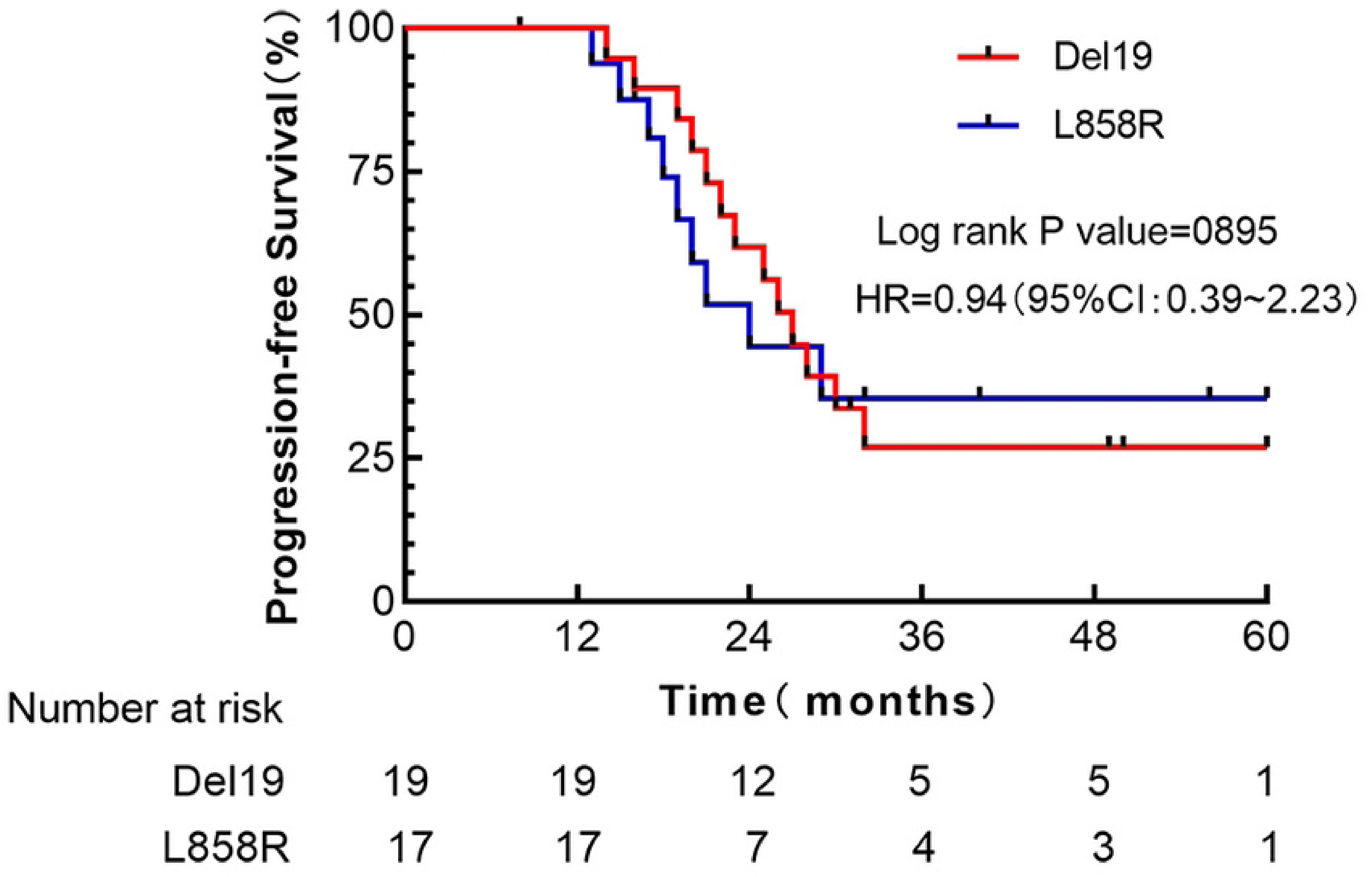
Kaplan-Meier curves of progression-free survival for exon 19 deletion mutation and exon 21L858R in the RT+TKI group(n=36). *Abbreviations:* CI=confidenceinterval; HR=hazard ratio.

**Fig 4B.**
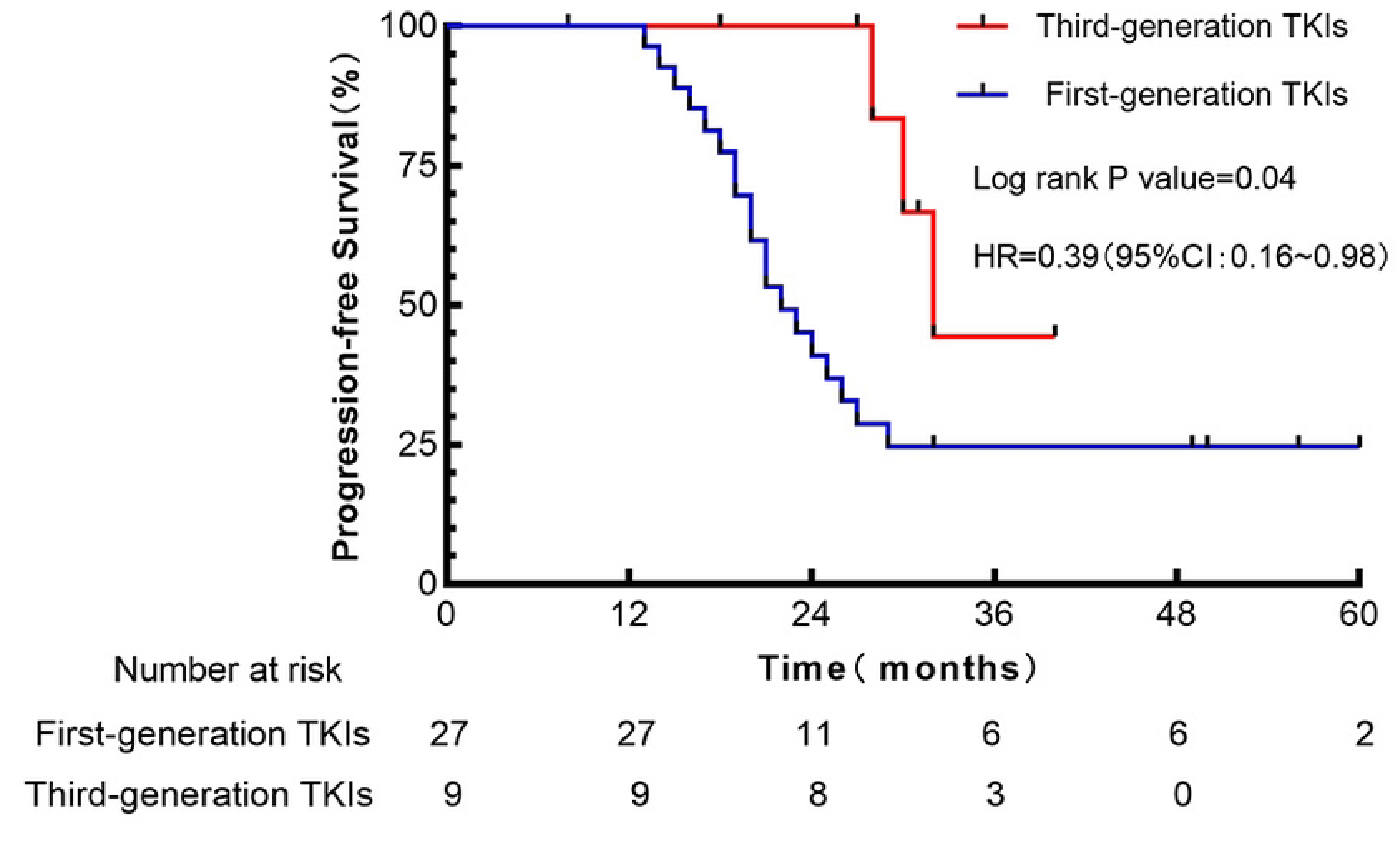
Kaplan-Meier curves of progression-free survival for Third and First-generation TKIs in the RT+TKI group (n=36). *Abbreviations:* CI=confidenceinterval; HR=hazard ratio.

**Fig 4C.**
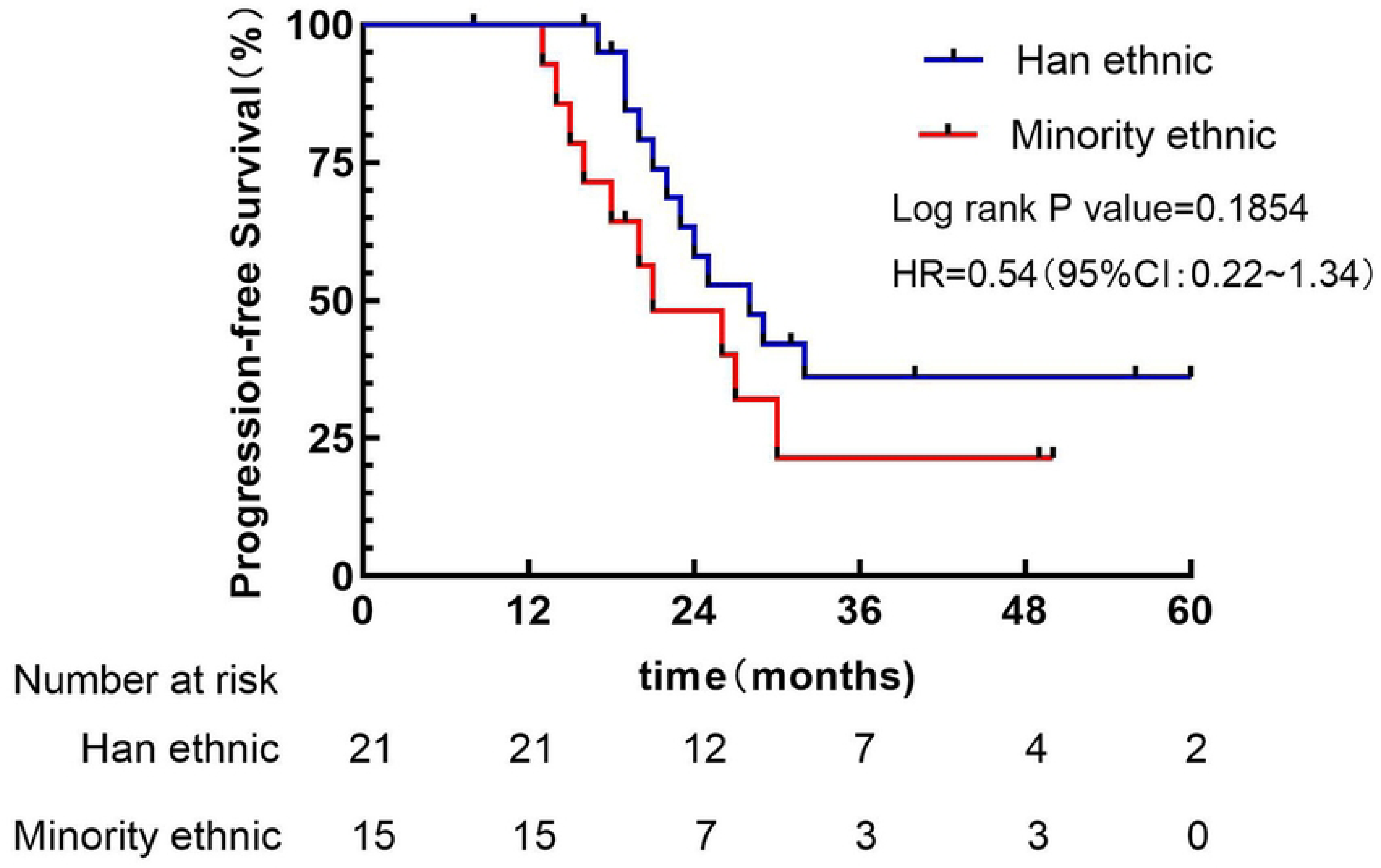
Kaplan-Meier curves of progression-free survival for Minority and Han ethnic in the RT+TKI group(n=36). *Abbreviations:* CI=confidenceinterval; HR=hazard ratio.

The 3 and 5-year OS rates for the RT+TKI group were 69.9 and 51.3%, respectively. The RT+CT group had 3 and 5-year OS rates of 67.3 and 43.6%, respectively. The difference in OS rates between the two groups was not statistically significant (χ2=0.049, P=0.825)(Fig 5.). Subgroup analysis revealed that there were no significant differences in OS between RT+TKI and RT+CT, regardless of age (≥ 60 or <60 years), sex (male or female), smoking status, ethnicity (Han Chinese or minority) or presence of Del19 or L858R mutations(Fig 6.).

**Fig 5.**
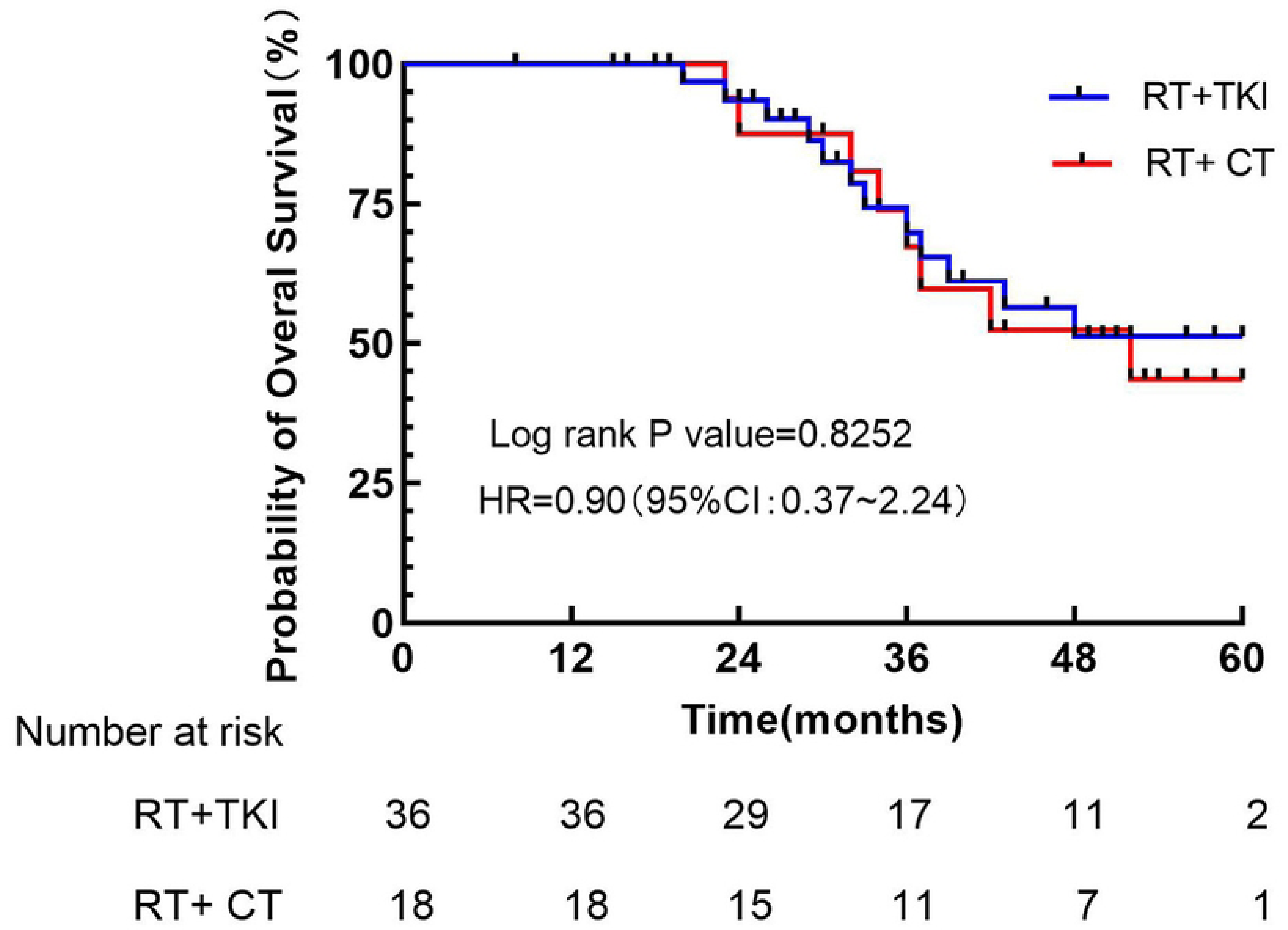
Kaplan-Meier curves of overal survival for all patients(N=54). *Abbreviations:* CI=confidenceinterval; HR=hazard ratio; RT=radiotherapy; TKI=tyrosine kinase inhibitors; CT=chemotherapy.

**Fig 6.**
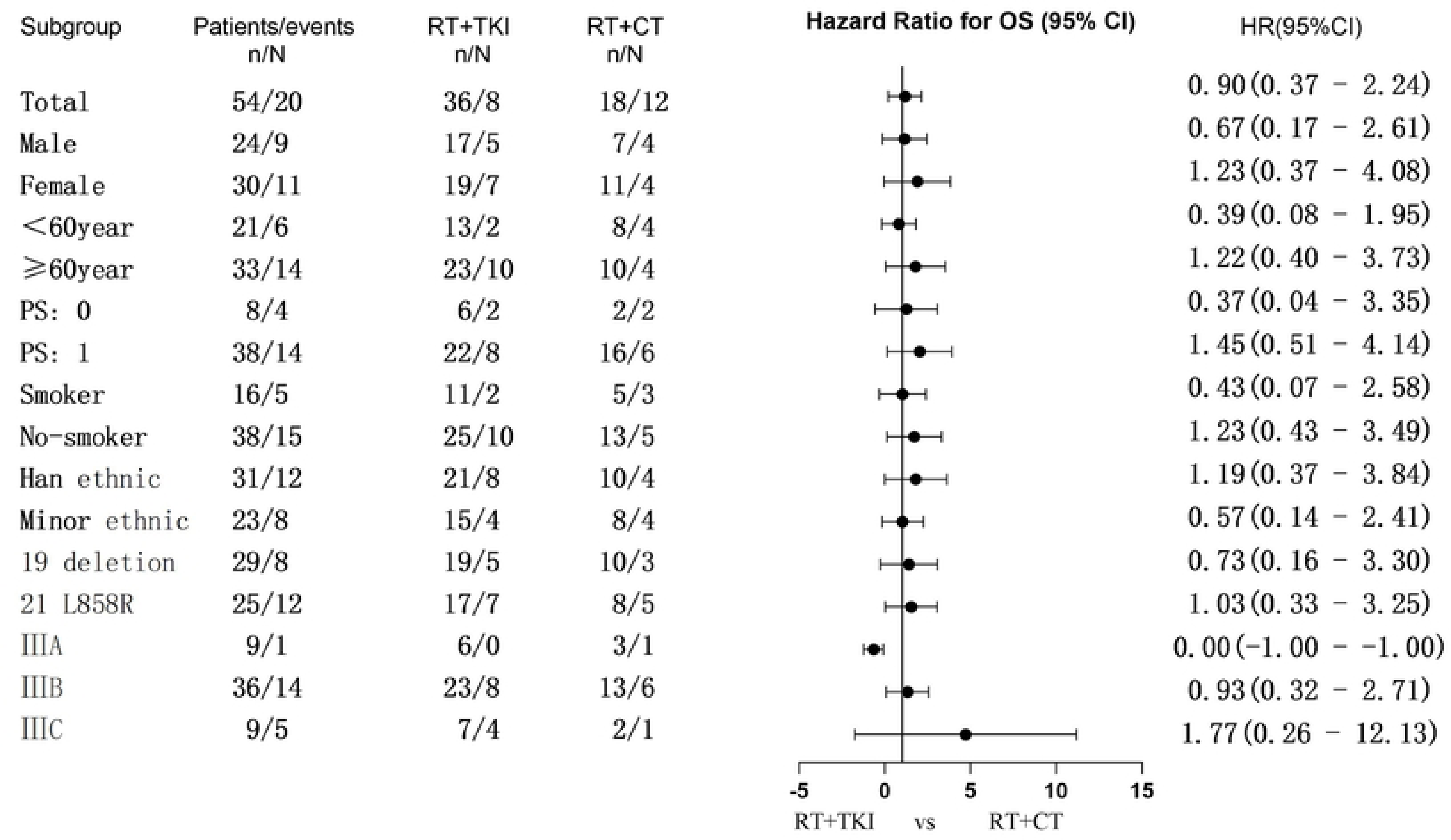
Forest plots for evaluation of treatment effect on different group variable. The analysis was performed on 54 patients with EGFR mutations with OS as the endpoint. *Abbreviations:* CI=confidenceinterval; HR=hazard ratio; PS=Performance Status; RT=radiotherapy; TKI=tyrosine kinase inhibitors; CT=chemotherapy.

### Patterns of Treatment Failure

At the time of data cut-off, treatment failure was observed in 22 out of the 36 patients (79.2%) in the RT+TKI group, which was primarily due to the fact that the cases that did not achieve CR underwent tumor progression. Among these, 4 cases were characterized by sole progression of the primary tumor, 15 cases exhibited sole appearance of new lesions, and 3 cases involved both progression of the primary lesion and emergence of new lesions. The most common site for the appearance of new lesions was intracranial (6 cases), followed by lung (4 cases), bone (3 cases), lymph nodes (3 cases)and liver (2 cases). In the RT+CT group, 16 cases experienced tumor progression, including 5 cases with the sole progression of the primary tumor, 8 cases with sole appearance of new lesions, and 3 cases with both the progression of the primary lesion and emergence of new lesions(Table IV).

In the RT+TKI group, among the 22 patients with tumor progression, 19 received salvage treatment, including switching to third-generation TKIs, stereotactic body radiotherapy (SBRT), particle implantation, brain RT, systemic chemotherapy and immunotherapy. In the RT+CT group, all 16 patients with tumor progression received salvage treatment, mainly transitioning to TKIs therapy, with few patients also undergoing subsequent SBRT.

### Adverse reactions

In terms of adverse reactions, the RT+TKI group exhibited milder systemic reactions, all graded at levels 1-2. By contrast, the RT+CT group experienced more severe systemic reactions, particularly in gastrointestinal and hematologic toxicities. The incidence of grade ≥3 gastrointestinal toxicity reached 38.9%, while grade ≥3 hematologic toxicity reached 33.3%. Compared with the RT+TKI group, these differences were statistically significant. Both the RT+TKI and RT+CT groups encountered varying degrees of radiation-related pneumonitis and esophagitis. The incidence of grade ≥3 radiation pneumonitis was 22.2% in the RT+TKI group and 27.8% in the RT+CT group, with no statistically significant difference (χ2=0.120, P=0.729). One case of grade 4 radiation pneumonitis occurred in the RT+TKI group, involving a patient with T2N3 peripheral lung cancer who received a prescribed dose of 66 Gy in 33 fractions, with lung Dmean of 16.29 Gy and V20 of 29.9%(Fig 7A.).

**Fig 7A.**
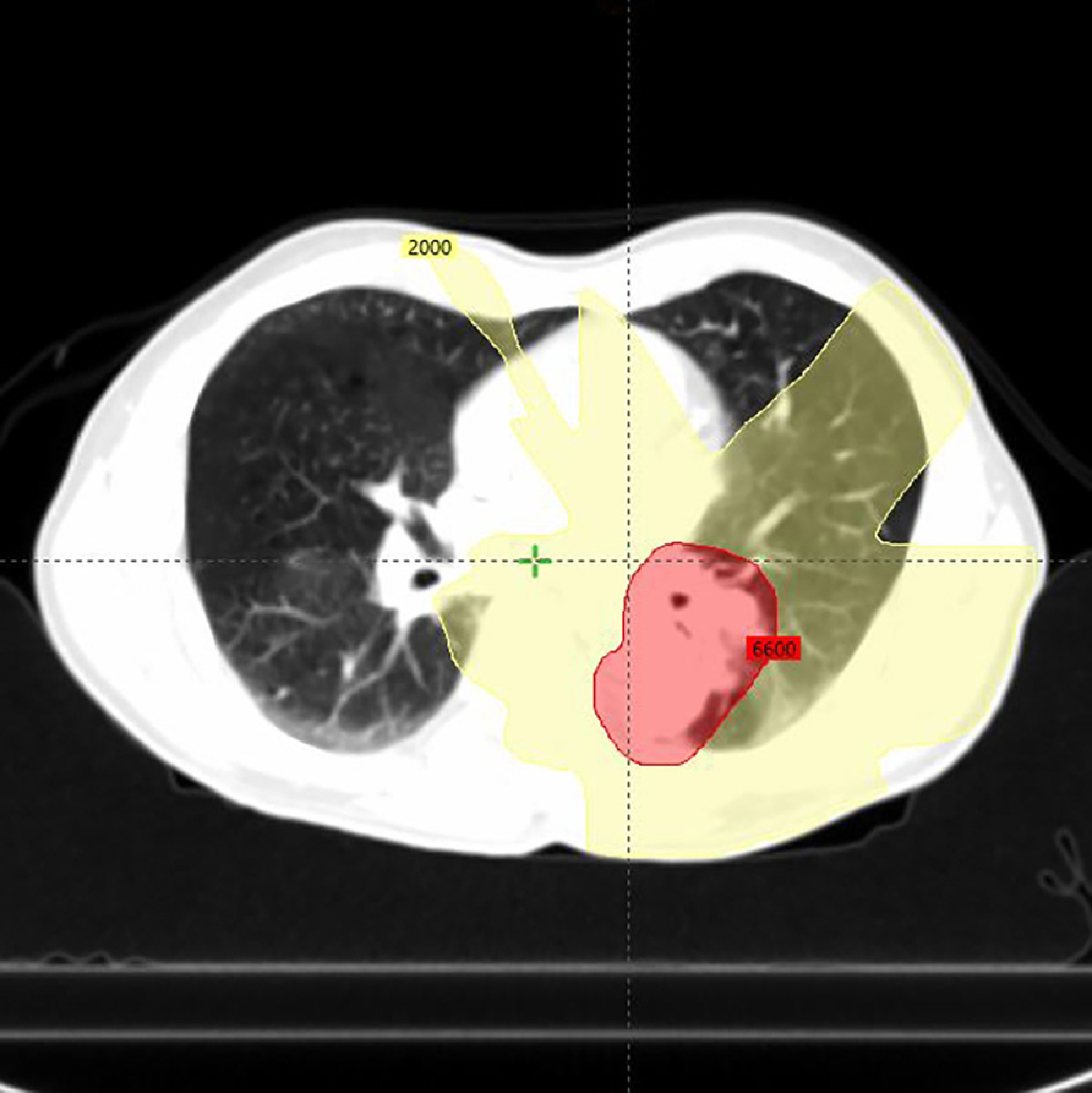
The dose distribution curve for the patient who experienced Grade 4 radiation-induced lung injury.

The TKI used in this case was gefitinib. Unfortunately, on the second day after completing 66 Gy/33 fractions, the patient experienced severe dyspnea, making it difficult to lie flat and requiring oxygen support. After receiving steroid pulse therapy, the patient’s symptoms gradually improved. One month after radiotherapy, a follow-up chest CT revealed CR of the tumor, but there was still some inflammation in the irradiated lung(Fig 7B.). In the DVH plot, it can be observed that V20 is <30%, but V5 is relatively high, reaching almost 70%(Fig.7C.). No grade 5 adverse reactions were reported in the RT+TKI or RT+CT groups(Table V).

**Fig 7B.**
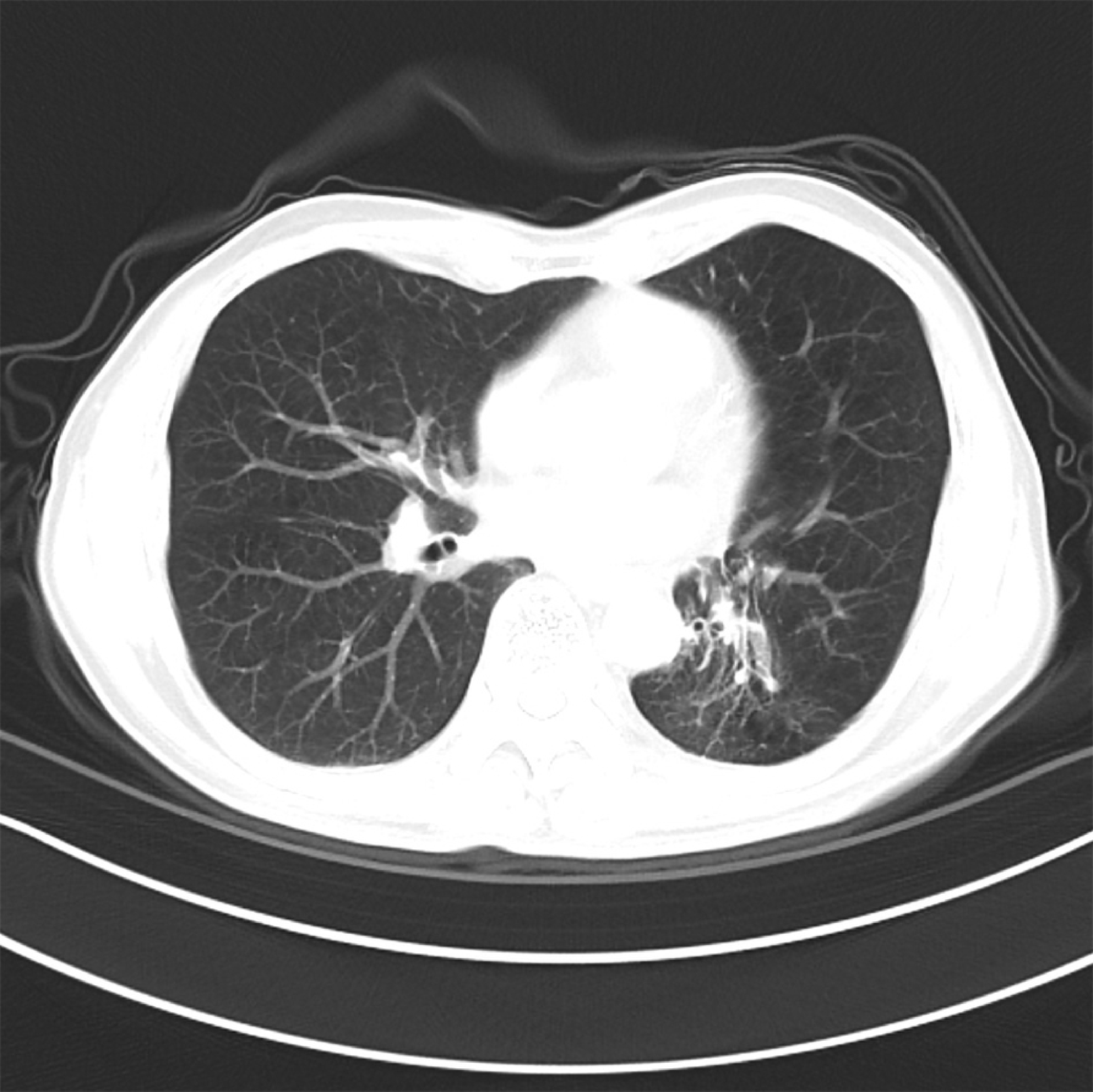
The CT image taken one month after radiotherapy for the patient who experienced Grade 4 radiation injury shows complete resolution of the tumor, but inflammation, infiltration, and fibrosis are observed in the lungs.

**Fig 7C.**
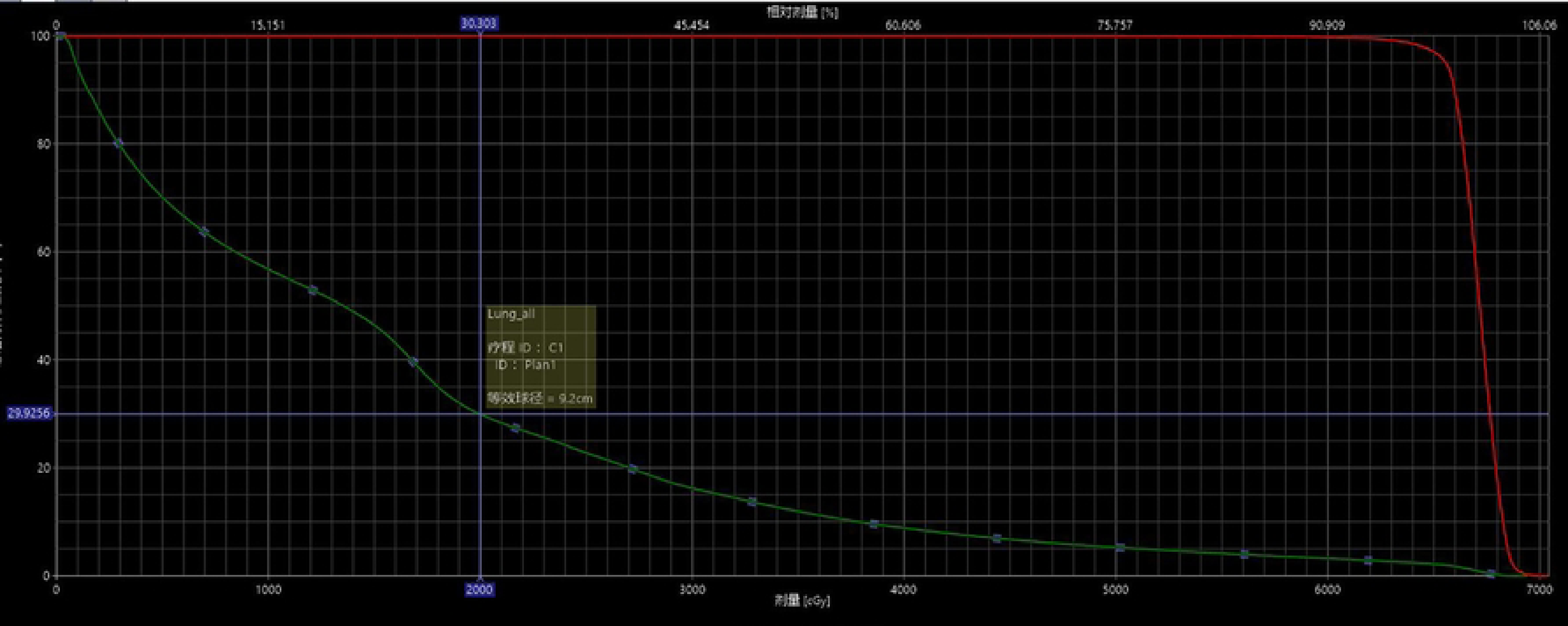
The DVH (Dose-Volume Histogram) plot for the patient with Grade 4 radiation injury.

## Discussion

The present study found no significant differences in short-term efficacy between RT with EGFR-TKIs and concurrent chemoradiotherapy for stage III EGFR-mutant lung cancer. However, in the long term, RT with EGFR-TKIs led to significantly prolonged PFS compared with concurrent chemoradiotherapy. This indicates that in real-world clinical practice, the efficacy of thoracic RT combined with EGFR-TKIs treatment for stage III EGFR-mutant lung cancer is superior to concurrent chemoradiotherapy.

In a phase II clinical study conducted in China (RECEL, NCT01714908) (18), RT combined with erlotinib was employed for the treatment of stage III unresectable EGFR-mutated NSCLC. The results indicated that the median PFS in the RT combined with erlotinib group was 24.5 months, compared with 9.0 months in the control group (concurrent chemoradiotherapy), demonstrating a statistically significant difference. Another phase II single-arm study from Japan (WJOG6911) (19), involving 27 cases of locally progressive unresectable EGFR-sensitive mutated NSCLC treated with RT combined with gefitinib, reported a median PFS of 18.6 months and a median OS of 61.1 months. A retrospective study conducted at the Cancer Hospital of the Chinese Academy of Medical Sciences, which included 511 patients with locally advanced EGFR-mutated lung adenocarcinoma, demonstrated median PFS values of 24.8, 12.6, and 15.9 months in the RT+TKI, CRT, and TKI groups, respectively, with statistically significant differences(20). The PFS outcomes from the present study were generally consistent with these previous studies, suggesting that RT combined with TKIs offers superior survival benefits compared to concurrent chemoradiotherapy for stage III EGFR-sensitive mutated lung cancer.

Notably, the PFS observed in the present study was prolonged more than that reported in previous studies(18–21), potentially due to the inclusion of cases involving third-generation TKIs, which had shown significantly longer OS in stage IV EGFR-sensitive mutated lung cancer(22). Subgroup analysis conducted in the RT+TKI group demonstrated that the median PFS was 32 months for RT combined with third-generation TKIs, whereas the median PFS for RT combined with first-generation TKI was 22 months, revealing a statistically significant difference (P=0.046). This finding suggests that the combination of RT and third-generation TKIs is more advantageous and superior in the treatment of stage III EGFR-mutated lung cancer compared with the combination of RT with first-generation TKIs. The primary reason for this disparity may be attributed to the ability of third-generation TKIs to effectively inhibit tyrosine kinase activation resulting from EGFR mutation and to overcome drug resistance caused by T790M mutation(23). It has been demonstrated that third-generation TKIs exhibit improved penetration through the blood-brain barrier, and in patients without brain metastases, it can also prolong the time until the development of brain metastases(24). It seems that the combination of RT with third-generation TKIs represents a preferable therapeutic option for stage III inoperable EGFR-sensitive mutated lung cancer. However, the third-generation TKI population is only 9 patients with shorter follow-up.This is attributed to the fact that the third-generation targeted therapy, specifically Osimertinib, was incorporated into first-line treatment indications in China only in 2019. Therefore, The conclusions drawn from this subgroup warrant a cautious interpretation.

Another subgroup analysis conducted in the RT+TKI group revealed that the disparity in PFS between the Del19 and L858R groups did not exhibit statistical significance, which contrasts with findings from previous studies. Earlier meta-analyses conducted in China confirmed that Del19 mutations were associated with longer PFS and OS compared with L858R mutations(25, 26). Subgroup analyses from the FLAURA and ACHEER1050 studies also demonstrated that the therapeutic benefit of TKI treatment in the L858R group was not as pronounced as that observed in the Del19 group(22, 27). In the present study, the median PFS was 27 months in the Del19 group and 24 months in the L858R group, suggesting a trend towards prolonged PFS in the Del19 group; however, the difference did not reach statistical significance, likely due to the limited sample size. This observation warrants further validation in future large-scale clinical trials.

Of note, the difference in the 3 and 5-year OS rates between the RT+TKI and RT+CT groups did not reach statistical significance, regardless of age (≥60 or <60 years), sex (male or female), smoking status, ethnicity (Han Chinese or minority), or the presence of Del19 or L858R mutations.This is primarily due to the fact that the majority of patients in both groups underwent salvage treatment after initial tumor progression. In the RT+CT group, patients primarily transitioned to TKI therapy, while in the RT+TKI group, the treatment was upgraded to include third-generation TKIs or shifted to chemotherapy, immunotherapy, and SBRT for localized progression, depending on individual conditions. Throughout this process, EGFR-TKIs played a pivotal role in both groups, underscoring the dominance of TKIs in the treatment of EGFR-mutant lung cancer. It is worth emphasizing that the results of the PACIFIC study indicated that patients with unresectable stage III EGFR-mutant lung cancer did not benefit from immunotherapy(28). Conversely, patients receiving TKIs treatment after chemoradiotherapy exhibited significantly higher disease-free survival rates than those receiving durvalumab(29). The results of the present study are consistent with the above findings, thus confirming the effectiveness of thoracic RT combined with TKI for the treatment of stage III EGFR-mutant lung cancer.

In terms of adverse reactions, the incidence of grade ≥3 hematological toxicity and gastrointestinal reactions in the RT plus targeted therapy group was significantly lower than that in the RT plus chemotherapy group. The occurrence rates of radiation-related pneumonia and esophagitis were comparable to those in the RT plus chemotherapy group, and the difference between the two was not statistically significant. This suggests that the safety profile of RT combined with EGFR-TKI is favorable. Radiation pulmonary toxicity remains a matter of concern in the context of combined RT and targeted therapy. A previous study have found that gefitinib could induce the occurrence of interstitial pneumonia, and when RT was performed on the basis of interstitial pneumonia, the severity of radiation lung injury was exacerbated(16). In the current study, the incidence of grade ≥3 radiation-associated pneumonia in the RT+TKI group was 22.2%, which, compared with that reported in previous studies, was not significantly increased. There was one case of grade 4 radiation-related pneumonia in a patient treated with RT combined with gefitinib, who had T2N3 peripheral lung cancer and was treated with a prescription dose of 66 Gy/33 fractions. The Dmean was 16.29 Gy, and V20 was 29.9%. A review of the imaging data for this patient did not reveal interstitial pneumonia outside the radiation field. Thus, it could not be conclusively determined whether the exacerbation of radiation-related lung injury was due to the use of gefitinib. Peled et al.(30) utilized neoadjuvant osimertinib treatment for inoperable stage III EGFR-mutant NSCLC, demonstrating a reduction of ≤50% in radiation field. Therefore, for lung cancer with larger tumors and broader radiation fields, administering induction TKI before RT appears to be a straightforward and effective approach.

There are two limitations to consider in the present study. Firstly, it is important to acknowledge that the current study design is retrospective, encompassing a relatively small sample size, which introduces the possibility of inherent bias. Therefore, caution must be exercised when drawing conclusions from the present findings. Secondly, the study period spans a considerable length of time, during which there have been rapid advancements in medical technology. Consequently, the results of the present study should be viewed primarily as a reference for clinical practice, considering the evolving landscape of medical advancements.

In conclusion, the combination of RT and TKIs exhibits superior efficacy and represents a safer therapeutic approach for the treatment of stage III EGFR-mutated lung cancer compared with concurrent radiochemotherapy, and such treatment modality may be regarded as a viable alternative in clinical practice.

## Data Availability

All relevant data are within the manuscript and its Supporting Information files.

## Acknowledgements

The authors would like to thank Professor Xiangpan Li (Oncology Center, The People’s Hospital of Wuhan University, Wuhan, China) and Dr Chunqiang Liu (The People’s Hospital of Laibin, Laibin, China) for their suggestions and help in writing the manuscript.

## Availability of data and materials

The data generated in the present study may be requested from the corresponding author, upon reasonable request.

## Authors’ contributions

LG: Conceptualization, methodology and writing of the original draft. WY: Data curation, formal analysis and software. GM: Visualization and investigation. ZQ: Supervision. HJ: Project administration. GL: Writing, reviewing and editing of the article.

## Ethics approval

The present study was conducted according to the Declaration of Helsinki and received approval from the Ethics Committee of The People’s Hospital of Laibin (approval no.2022-54.Laibin,China), The First People’s Hospital of Yulin (approval no. 2022-95. Yulin,China), and Guangxi Medical University Kailuan Langdong Hospital(approval no.2022-73.Nanning,China).

## Patient consent for publication

Not applicable

